# Anti-SARS-CoV-2 IgM and IgG antibodies in health workers in Sergipe, Brazil

**DOI:** 10.1101/2020.09.24.20200873

**Authors:** Mônica Santos de Melo, Lysandro Pinto Borges, Daniela Raguer Valadão de Souza, Aline Fagundes Martins, José Melquiades de Rezende Neto, Anderson Alves Ribeiro, Aryanne Araujo Santos, Grazielly Bispo da Invenção, Igor Leonardo Santos Matos, Kezia Alves dos Santos, Nicolas Alessandro Alves Souza, Pâmela Chaves Borges, Makson Gleydson Brito de Oliveira

## Abstract

**Background:** The exponential growth of COVID-19 cases in Brazil is overloading health systems with overcrowding of hospitals and overflowing intensive care units. Increasing infection rates in health professionals can lead to the collapse of the health system and further worsen the pandemic. The aim of this study was to evaluate the seroprevalence of IgM and IgG for SARS-CoV-2 in health workers in Sergipe, Brazil.

**Methods:** The targeted tests involved health professionals working on the front line to combat COVID-19. The samples were collected in the month of June, in six hospital units in the state of Sergipe.

**Results:** 471 health professionals were tested. Of these, 28 workers (5.95%) tested positive for IgM and 64 (13.59%) tested positive for IgG. 9 workers (1.91%) tested positive for IgM and were also positive for IgG.

**Discussion:** Health workers must be monitored constantly, because if they are infected, they can spread the virus to colleagues, hospitalized patients and even family members.

**Conclusion:** Knowing the prevalence of antibodies to the virus in health workers is an important measure of viral spread control.

## 1. Introduction

The SARS-CoV-2 infection started in Wuhan, spread further in China and then spread rapidly around the world, classified by the World Health Organization (WHO) on March 11 as a pandemic. This infection causes a severe acute respiratory syndrome, now called COVID-19 (HUANG *et al*., 2020; ZHU *et al*., 2020; WHO, 2020).

In addition to respiratory symptoms, gastrointestinal symptoms, impairment of the nervous system and a variety of clinical signs such as fever, cough, shortness of breath, sore throat, malaise too can be seen (SINGHAL, 2020; BENVENUTO *et al*., 2020). Other symptoms include headache, dyspnoea, hemoptysis, diarrhea and lymphopenia (ROTHAN and BYRAREDDY, 2020).

Infection with SARS-CoV-2 can be aggressive and generates a high rate of hospitalization and, in some cases, hospitalization in Intensive Care Units (BAUD *et al*., 2020). In Brazil, the exponential growth of COVID-19 cases is overloading health systems with overcrowding of hospitals and overflowing intensive care units. The Unified Health System (SUS) follows the guidelines recommended by the WHO to limit viral spread, isolating people who contract the virus and oriented to compliance with quarantine for people who are at greatest risk (OLIVEIRA *et al*., 2020).

The orientation of social isolation is not suitable for health workers who work directly to confront COVID-19. Increased infection rates among health workers can lead to the collapse of the health system and further worsen the pandemic (BARRANCO and VENTURA, 2020). In this context, the objective of this study was to evaluate the seroprevalence of IgM and IgG for SARS-CoV-2 in health workers, from different professions, in some health units in the state of Sergipe, Brazil.

## 2. Methods

The targeted tests involved health professionals working on the front line to combat COVID-19. The samples were collected in the month of June, in six hospital units in the state of Sergipe, in the municipalities of Aracaju, Nossa Senhora do Socorro, Nossa Senhora da Glória and Estância. Personal data including age, gender, address, presence of any comorbidities and symptoms compatible with COVID-19 were gathered through an online questionnaire performed by a health care worker just before blood sample collection. Venous blood samples were collected and centrifuged for serum separation. 471 people were enrolled in this investigation.

The sample chosen for testing for IgG and IgM antibodies to SARS-CoV-2 was human serum. For this purpose, approximately 3 mL of blood was collected from each volunteer. Venous puncture was the chosen technique, with two possibilities for collection sites: the antecubital fossa of the arm or the back of the hand, according to the volunteer’s preference. This sample was stored in a refrigerated environment to maintain viable biological properties for analysis.

At the laboratory, the whole blood sample was centrifuged to separate the serum from the other blood components. The centrifuge was programmed to perform 4000 rotations every 1 minute, for 10 minutes. The serum was packed in Eppendorf tubes and kept refrigerated at −20°C until the moment of use. This biological material was analyzed using the ichroma™ COVID-19 Ab test (Boditech Med Inc. Chuncheon, Korea) following the manufacturer’s instructions for the detection IgG and IgM antibodies to SARS-CoV-2 by the fluorescence method.

Immunofluorescence assays were performed at the Department of Pharmacy Laboratory (Laboratory of Biochemistry and Clinical Immunology, LaBiC-Imm) at the Federal University of Sergipe (UFS). Anti-SARS-CoV-2 IgM and IgG antibodies were detected in sera using an in vitro diagnostic test system based on lateral flow sandwich detection immunofluorescence technology (Ichroma2™ COVID-19 Ab in conjunction with an Ichroma™ II Reader, Boditech Med Inc., South Korea) according to the manufacturer’s instructions. The immunofluorescence method applied showed a sensitivity of 95.8% and a specificity of 97%. Another work by our group carried out the validation of the method using 120 serum samples collected from 60 real-time reverse transcription polymerase chain reaction (rRT-PCR) confirmed COVID-19 cases, and 60 negative patients at different clinical sites (BORGES *et al*., 2020).

## 3. Results

A total of 471 health workers were tested. Of these, 134 were from the Sergipe Emergency Hospital, 33 from the University Hospital and 88 from the Nossa Senhora de Lourdes Maternity, all in the city of Aracaju; 76 of the Regional Hospital of Nossa Senhora da Glória; 66 of the Municipal Hospital of the city of Nossa Senhora do Socorro; 74 of the Regional Hospital of the city of Estância. Of these, 28 workers (5.95%) tested positive for IgM and 64 (13.59%) tested positive for IgG. These results indicate an active or recent infection for IgM reagents and a past infection for IgG reagents. 9 workers (1.91%) serological profile indicating a recent infection that can still be contagious, tested positive for IgM and were also positive for IgG. These data are plotted in Table 1. 370 workers tested non-reactants for the presence of IgM and IgG antibodies to SARS-CoV-2.

**Table 1:** Seroprevalence of IgM and IgG for SARS-CoV-2 in health workers in six hospital units Sergipe, Brazil

| Site | Valid samples | IgM reagent | IgG reagent | IgM and IgG reagent | No reagent |
| --- | --- | --- | --- | --- | --- |
| SEH | 134 | 1 | 13 | 1 | 119 |
| NSLM | 88 | 8 | 5 | 0 | 75 |
| RHNSG | 76 | 4 | 4 | 3 | 65 |
| MHNSS | 66 | 9 | 12 | 5 | 40 |
| UH | 33 | 2 | 7 | 0 | 24 |
| RHE | 74 | 4 | 23 | 0 | 47 |
| **Total** | **471** | **28** | **64** | **9** | **370** |
| **Total (%)** | **100%** | **5,95%** | **13.59%** | **1,91%** | **78,55%** |

**SEH:** Sergipe Emergency Hospital; **NSLM:** Nossa Senhora de Lourdes Maternity; **RHNSG:** Regional Hospital of Nossa Senhora da Glória; **MHNSS:** Municipal Hospital of Nossa Senhora do Socorro; **UH:** University Hospital; **RHE:** Regional Hospital of Estância.

## 4. Discussion

The approach to screening health professionals who participated in the study in the health environment depended on the institution’s policies, including the number of samples for each sector. In general, healthcare professionals should monitor themselves for fever and symptoms of COVID-19 and stay home if they are ill. In a report of 48 health workers with confirmed COVID-19 in King County, Washington, 65 percent reported working for an average of two days while exhibiting symptoms of COVID-19 (CHOW *et al*., 2020). In addition, symptom screening alone did not identify all cases. The survey aims to contribute to map the characteristics related to the work process of these professionals in the midst of the COVID-19 pandemic, as well as the initiatives that are being carried out by managers in health establishments in Brazil.

Based on these data, we can suggest the adoption of safety measures by health professionals in their work processes, focusing mainly on the proper use of Personal Protective Equipment (PPE); as well as Organization of the health establishment in relation to the supply of equipment and supplies, cleaning and disinfection of the environments, in addition to the training and monitoring indicators of its employees (LARSON and LIVERMAN, 2011).

The rapid and accurate diagnosis of COVID-19 contributes to the management of the disease (JIN *et al*., 2020). The identification of antibodies, mainly IgM and IgG immunoglobulins, produced shortly after infection, can contribute to ensure clinical diagnosis and monitoring of the disease, allowing early intervention and playing a key role in fighting outbreaks (BROADHURST *et al*., 2016; KLUGE *et al*., 2018). Among the serological tests available are ELISA, CLIA, chemiluminescence, fluorescence, qualitative immunochromatography (PADOAN *et al*., 2020; LIPPI *et al*., 2020; KIM *et al*., 2020; DI MAURO *et al*., 2020; CARVALHO *et al*., 2020). The immunofluorescence method was chosen for serological tests performed in this study. Anti-SARS-CoV-2 IgM and IgG antibodies were detected in the serum using an in vitro diagnostic test system based on lateral flow sandwich detection immunofluorescence technology, which presented a sensitivity of 95.8% and specificity of 97%.

Our results show 5.95% IgM reagent antibody, so the choice of serum as a biological sample for evaluation was due to the presence of antibodies. The production of antibodies, mainly IgM, produced quickly after infection, can be a useful tool to assist in the detection of contact with SARS-CoV-2. Thus, using laboratory techniques, the IgM - IgG antibody test allows early intervention with infected people, playing a critical role in fighting outbreaks (BROADHURST *et al*., 2016; KLUGE *et al*., 2018). Consistent with the recommendations of the Infectious Diseases Society of America (IDSA), to maximize the predictive value of the serological test, tests with high specificity (≥99.5 percent) should be used and the test should be reserved for individuals with high probability pretest of previous infection (HANSEN *et al*., 2020). Our group recently published a cross-sectional study with stratified sampling on the prevalence of SARS-CoV-2 antibodies in an asymptomatic population in the state of Sergipe. Samples from 3.046 asymptomatic individuals showed a high prevalence of SARS-CoV-2 antibodies (BORGES *et al*., 2020).

In contrast to data in the literature, detectable antibodies generally take several days to weeks to develop, and the time to detect antibodies varies according to the test (GUO *et al*., 2020; ZHAO *et al*., 2020; QU *et al*., 2020; ZHANG *et al*., 2020). For example, in a study that considers 38 studies in a systematic review, IgM was detected in 23 percent in one week, 58 percent in two weeks and 75 percent in three weeks; the corresponding detection rates for IgG were 30, 66 and 88 percent (DEEKS *et al*., 2020). Other studies have suggested that the positive IgG rate approaches 100 percent in 16 to 20 days (CATUREGLI *et al*., 2020; LONG *et al*., 2020a; WANG *et al*., 2020). According to our data, 13.59% of reagent IgG is observed. However, the duration of the detectable antibodies is uncertain (LONG *et al*., 2020b; IBARRONDO *et al*., 2020). In one study, it was observed that IgG levels decreased by an average of approximately 75 percent from the acute to the early convalescent phase of the disease and, eight weeks after infection, 40 percent of asymptomatic patients and 13 percent of symptomatic patients had no detectable IgG (LONG *et al*., 2020b).

Although people should stay at home to reduce the spread of COVID-19, health professionals do the just the opposite. Your job and the longest job hours (due to the increase in the number of infected people hospital) put them at risk of infection (THE LANCET, 2020). Health professionals must be monitored constantly, because if they are infected, they can spread the virus to colleagues, hospitalized patients and even family members. Increased infection rates in healthcare professionals can cause the pandemic to worsen, so an adequate supply of effective PEPs, careful monitoring of all healthcare professionals is essential (BARRANCO and VENTURA, 2020). This is due to the fact that health professionals are currently the most important resource and, therefore, if health professionals are not cared for, the health system is severely damaged.

## 4. Conclusion

The study showed the prevalence of COVID-19 in health professionals in the state of Sergipe, Brazil. The immunofluorescence assay revealed the antibody rate for SARS-CoV-2 in the studied population. Knowing the prevalence of antibodies to the virus in health workers is an important measure of viral spread control, since they do not comply with the recommendations of social isolation, due to the need to act in the time of a pandemic, and, therefore, are most susceptible to viral contamination and spread.

## Data Availability

I declare that all data published in this study are available for consultation.

## Author Contributions

All authors have read and agreed to the published version of the manuscript.

## Funding

This research was not funded. The authors themselves financed it.

### Acknowledgments

The authors dedicate this article to all health professionals who are facing COVID-19. We are eternally grateful to them, and hope that our article can contribute to reducing the number of deaths.

